# Clinical Validation of Automated and Rapid mariPOC SARS-CoV-2 Antigen Test

**DOI:** 10.1101/2021.02.08.21250086

**Authors:** Juha M. Koskinen, Petri Antikainen, Kristina Hotakainen, Anu Haveri, Niina Ikonen, Carita Savolainen-Kopra, Kati Sundström, Janne O. Koskinen

## Abstract

Novel SARS coronavirus causing COVID-19 was recognized in late 2019. Diagnostics was quickly ramped up worldwide based on the detection of viral RNA. Based on the scientific knowledge for pre-existing coronaviruses, it was expected that the RNA of this novel coronavirus will be detected from symptomatic and at significant rates also from asymptomatic individuals due to persistence of non-infectious RNA. To increase the efficacy of diagnostics, surveillance, screening and pandemic control, rapid methods, such as antigen tests, are needed for decentralized testing and to assess infectiousness. The objective was to validate the analytical and clinical performance, and usability of a novel automated mariPOC SARS-CoV-2 test, which is based on the detection of structural viral proteins using sophisticated optical laser technology. Clinical performance of the test was evaluated against qRT-PCR with nasopharyngeal swab specimens collected from patients suspected of acute SARS-CoV-2 infection. Sensitivity of the mariPOC test was 100.0% (13/13) directly from swab specimens and 84.4% (38/45) from swab specimens in undefined transport mediums. Specificity of the test was 100.0% (201/201). The test’s limit of detection was 2.7 TCID_50_/test and had no cross-reactions with the tested respiratory microbes. Our study shows that the mariPOC can detect infectious individuals already in 20 minutes with clinical sensitivity close to qRT-PCR. The test targets conserved epitopes of SARS-CoV-2 nucleoprotein, making it robust against strain variations. The new test is a promising and versatile tool for syndromic testing of symptomatic cases and for high capacity infection control screening.

## Introduction

Emerging pandemic coronavirus (CoV) was recognized in Wuhan, China, in late 2019. The virus, isolated from patients mentioned to be pneumonic, was quickly sequenced to share 79.6% full length genome similarity with the Severe Acute Respiratory Syndrome virus (SARS-CoV-1) and 91.2% similarity between its nucleocapsid (N) proteins (1). The novel SARS-CoV-2, causing COVID-19, was identified to be circulating in horseshoe bats for decades similarly to SARS-CoV-1 (2). Diagnostic nucleic acid amplification tests (NAAT), mostly quantitative real-time polymerase chain reaction (qRT-PCR), were quickly developed worldwide, based on protocol provided for World Health Organization (3). Diagnostic qRT-PCR capacities were ramped up quickly in central laboratories because such tests are fast to develop for new targets. Most often, the new qRT-PCR tests were adopted for clinical diagnostics with minimal verification and validation against other diagnostic test methods.

For the seasonal coronaviruses, the interpretation of gene positivity in clinical specimens has been challenging since the viral RNA is detected at similar rates and qRT-PCR cycle threshold (Ct) values from symptomatic and asymptomatic individuals. The viral RNA is also co-detected with genomes of other respiratory viruses (4-7). This is also the case for the SARS-CoV-2 (8, 9). Moreover, recent scientific evidence indicates that qRT-PCR positivity has poor correlation for assessment of SARS-CoV-2 infectiousness (10-16). Whereas, Pekosz et al. (2020) showed that the detection of N-protein by an antigen test correlates with SARS-CoV-2 viral culture more accurately than qRT-PCR (13). Already half a decade ago Inagaki et al. (2016) unequivocally concluded for influenza that, “PCR…is not an appropriate method for indicating infectivity” and “the antigen-detection test estimated the infectious period with comparable if not better accuracy with culture” (17). In the case of COVID-19 diagnostics, the fact that viral RNA persistence can be detected without viable virus for months, has been a known clinical challenge, as diagnostics relied in the beginning of the pandemic solely on NAAT detection (18), the efficacy of which is in ruling out positivity.

The expression of N-protein, which is the key pathogenicity factor of coronaviruses (19), is essential for the coronavirus replication and transcription of the viral RNA (20, 21). Without the accumulation of the N-protein, the coronaviral mRNA is degraded by the nonsense-mediated decay (NMD) pathway of eukaryotic cells (19). Alexandersen at al. (2020) concluded that the detection of RNA is not an indicator of actively replicating SARS-CoV-2. Their data suggests that virion and subgenomic RNAs are stable in cellular double-membrane vesicles and, therefore, can be detected long after the acute infection (22).

Shortly after viral exposure, viral concentration is low and qRT-PCR Ct values are high. When the virus starts replication, it happens fast. In a cell model, extensive coronavirus RNA transcription has occurred in 6 to 8 hours after the infection (23). In addition, NAATs being prone for reporting clinically insignificant findings (analytically the detection may be correct, there is viral RNA in the sample) they are prone to contaminations. A study of SARS-CoV-2 primer-probe sets from four major European suppliers found a significant level of contamination from the reagents. False positives as low as qRT-PCR Ct 17 were obtained (24). Low levels of SARS-CoV-2 RNA contamination has also been found from surfaces and air in rooms where mildly ill individuals were isolated without notable viable virus (25, 26). It has also been shown that environmental contamination may yield in positive test results in PCR among individuals sampled in the same area where intranasal influenza vaccine dosing was done (27). These data suggests that individuals having presence near symptomatic patients can be contaminated by RNA without being infected with viable virus. Thus, methods detecting the viral RNA by amplification are prone for clinically insignificant positive results, especially when significant part of the population has been infected recently. The fact that a positive NAAT result is not a reliable biomarker of active infection or COVID-19, is a true challenge for clinicians and decision making for quarantine. It is not only that a missed necessary quarantine has health and epidemic costs but also that a falsely imposed quarantine has social and financial consequences (28).

The different performance requirements of diagnostic, surveillance and screening testing have been recently discussed by Mina and Andersen (2020). There is a need for both super sensitive PCR based tests and rapid and appropriately sensitive antigen tests to fight the COVID-19 pandemic (29). The use of the two methodologies should supplement one another in clinical practice and pandemic fight.

In the present study, we analytically and clinically validated the performance of a novel 2nd generation mariPOC SARS-CoV-2 test (ArcDia International Ltd, Finland), intended for rapid and automated detection of viral acute phase proteins when there is a clinical suspicion of acute COVID-19. Monoclonal antibodies of the test are designed to target a conserved epitope in the N-protein, which is the most abundant protein in coronaviruses. We have previously shown that the presence of coronavirus OC43 N-protein in the nasopharynx correlates with the respiratory tract infection symptoms (30). It has been shown that clinical presentations of seasonal coronavirus OC43 infections can be similar to those of coronaviruses that are considered as severe viruses (SARS and MERS) (31).

The mariPOC is an automated platform for the rapid multianalyte testing of acute infectious diseases. It is based on a separation-free two-photon excitation assay technique (32, 33). Subsequently to nasopharyngeal sampling, the mariPOC test’s operational steps are: combining the swab with the sample buffer from a bottle-top dispenser, vortexing the sample tube to release the sample from the swab (Fig. 1), followed by automated analysis and objective fluorescent result read out. The analysis is performed by an automated analyzer with sophisticated autoverification functions, and the result can be transferred automatically to the laboratory information system or as anonymized epidemiological data (34) into mariCloud™ service. The hands-on time is one minute per sample, and the analyzer works in continuous-feed and walk-away mode. The mariPOC SARS-CoV-2 test is available as a single pathogen test and as part of syndromic multianalyte tests covering, among others, influenza viruses. The throughput of one mariPOC analyzer can be up to 300 single analyte tests or 100 multianalyte tests in 24 hours. The results are reported in two phases, great majority of the infectious cases in twenty minutes and very low positive and negative cases in 55 minutes or two hours, depending on the test configuration.

**Fig. 1.**
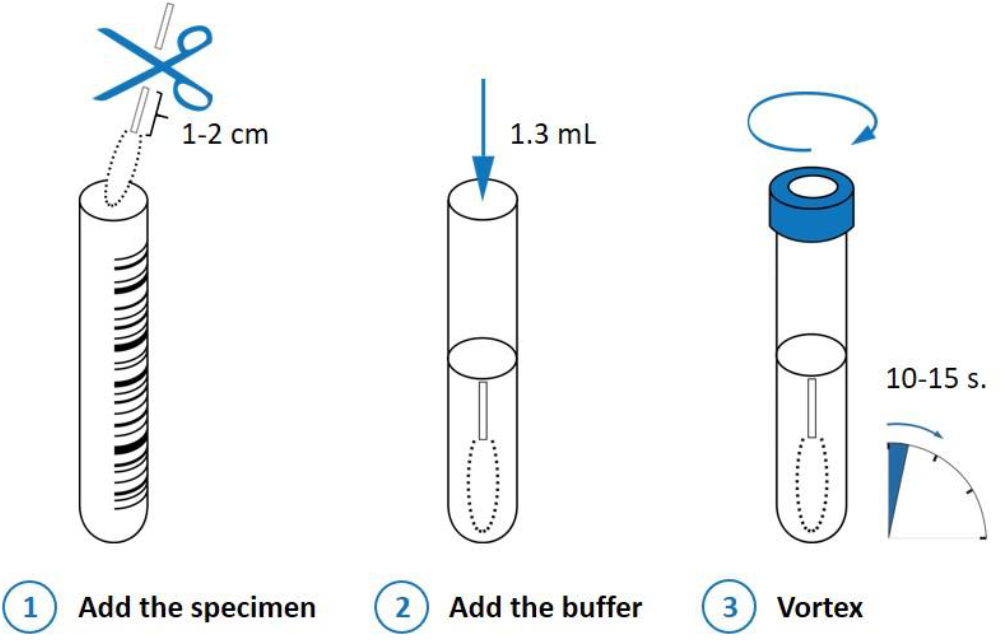
Sample pretreatment in the mariPOC SARS-CoV-2 test before placing the sample tube into the analyzer for automated analysis and subsequent test result read out objectively as positive or negative.

## Materials and methods

### Analytical sensitivity

Nasopharyngeal swab specimens from asymptomatic individuals were pooled and suspended into mariPOC RTI sample buffer into volume corresponding to 1.3 mL per swab. This pooled clinical sample matrix (1.3 mL) was spiked with 75 µL of gamma-irradiation inactivated SARS-CoV-2 cell lysate (stock 2.8 × 10^6^ TCID_50_/mL, USA-WA1/2020, NR-52287, lot 70035888, BEI Resources, Manassas, VA, USA) in culture supernatant of different viral concentrations (50% tissue culture infective dose, TCID_50_). The samples were analyzed with the mariPOC SARS-CoV-2 test following the manufacturer’s instructions. Limit of Detection (LoD) was determined as the lowest concentration giving at least 19 positives out of 20 replicates (≥95% positivity).

### Cross-reactivity

Analytical specificity of the mariPOC SARS-CoV-2 test was studied by challenging the test against relevant microbes commonly found in the nasal cavity (Online Resource). Briefly, the microbe stocks were suspended in high concentration in the mariPOC RTI sample buffer and analyzed with the mariPOC test.

### Clinical specificity

Validation of the specificity of the mariPOC SARS-CoV-2 test was conducted in SataDiag laboratory unit in Pori, Finland in February 2021 by one operator following the manufacturer’s instructions. In the study, 205 freshly sampled nasopharyngeal swab specimens were analyzed. The samples were leftover samples from routine diagnostics with the 1st generation (launched May 2020) mariPOC SARS-CoV-2 test. Samples positive in mariPOC were also analyzed with the Xpert Xpress SARS-CoV-2 test (ref XPRSARS-COV2-10) detecting N2 and E-genes, Cepheid, USA.

### Clinical sensitivity

Sensitivity of the mariPOC SARS-CoV-2 test was validated with 58 frozen qRT-PCR positive nasopharyngeal samples from two specimen cohorts.

#### Sample cohort 1

The first cohort consisted of 13 qRT-PCR positive nasopharyngeal swab samples collected from patients (N=211) visiting primary healthcare COVID-19 drive-in stations of Mehiläinen Oy in Helsinki capital area of Finland from March to April 2020. The qRT-PCR negative frozen samples were not analyzed for this study because the test specificity was studied with fresh samples as described above. The enrollment criteria were respiratory infection symptoms and clinician’s suspicion of COVID-19, the official criteria for COVID-19-testing in Finland, and at the clinical study sites at the time of the study. The samples were taken with a flocked swab from the nasopharynx (8 to 12 cm deep for adults and 4 to 8 cm deep for children) by rotating the swab in nasopharyngeal cavity for 10 seconds. Two consecutive specimens were collected from 127 patients giving oral consent to participate in the study. The specimens were collected during an internal laboratory method validation study, which does not require external ethical review board permission and was not linked with recruitment or treatment of patients (35). The study was approved by the responsible chief physician of Mehiläinen Oy, and by the responsible chief physician and area manager of SataDiag laboratory division. The specimen for standard of care testing was collected first. These specimens were analyzed after RNA extraction with Allplex™ 2019-nCoV RT-PCR assay (Seegene Inc., Republic of Korea) at Seoul Clinical laboratories (Republic of Korea). Allplex™ 2019-nCoV RT-PCR assay detects E, N and RdRP genes (36).

The second swab specimen was kept in a dry tube at +4 °C for a maximum of 8 hours and stored frozen until analysis with the mariPOC SARS-CoV-2 test. For 84 patients, one nasopharyngeal swab specimen was collected. These swabs were suspended into saline (0.5−1 mL) and analyzed with Amplidiag COVID-19 qRT-PCR assay including RNA extraction (Mobidiag Ltd, Finland) at Vita Laboratorio Ltd (Finland). Amplidiag COVID-19 qRT-PCR assay detects N and ORF1ab genes (37). The leftover saline specimens were stored frozen until analysis with the mariPOC SARS-CoV-2 test.

The dry swab specimens and leftover saline samples were analyzed retrospectively with the mariPOC test by two operators following the manufacturer’s instructions. Briefly, dry swabs were suspended into 1.3 mL of the mariPOC RTI sample buffer and the leftover saline samples (range 0.1−0.65 mL) were diluted with mariPOC RTI sample buffer to a final volume of 1.3 mL. For this validation study, the samples were further diluted into the mariPOC RTI sample buffer in one-to-one (1:1) ratio and analyzed with the mariPOC test. When a discrepant result between the mariPOC and comparator RT-PCR was obtained, the swab samples taken for the mariPOC were confirmatory tested at the Department of Clinical Microbiology, Turku University Hospital (Finland) with an in-house reference qRT-PCR detecting E, N and RdRP genes (3).

#### Sample cohort 2

The second cohort consisted of forty five positive pseudonymized specimens with known qRT-PCR Ct values (16 to 34) from the frozen nasopharyngeal swab specimen library of Finnish Institute of Health and Welfare, Helsinki, Finland, in undefined transport mediums. The qRT-PCR protocol was an in-house method based on the primers and probes by Corman et al. (2020) (3). The cohort consisted mostly of symptomatic, but in part also of asymptomatic subjects, while the detailed information for each subject was not available for this study. The specimens were in either reddish (N=22) or colorless (N=23) solutions. The samples were diluted into the mariPOC RTI sample buffer in one-to-one (1:1) ratio and analyzed with the mariPOC test. The positivity rate of the mariPOC test was evaluated against different qRT-PCR Ct value categories and then compared to published viral culture studies.

## Results

### Analytical sensitivity

LoD of the mariPOC SARS-CoV-2 test was 2.7 TCID_50_/test in 20 µL reaction volume, at which all twenty replicates gave a positive test result. Based on the certificate of analysis of the viral preparation, the LoD equals to 1690 genome equivalents per test.

### Cross-reactivity

The mariPOC test had no cross-reactions. The mariPOC test did not cross-react with seasonal coronaviruses OC43, 229E, or NL63, but it detected the recombinant N-protein of SARS-CoV-1. Detailed information is shown in supplemental material (Online Resource).

### Clinical specificity

Specificity of the mariPOC test was 100.0% in the preliminary (200/200) and final (201/201) result reporting phases. Three and two of the sample analyseswere rejected by the autoverification in the preliminary and final result reporting phases giving failure rates of 1.5% and 1.0%, respectively. Two of the samples were positive in both the mariPOC test and routine PCR test.

### Clinical sensitivity

The sensitivity of the mariPOC SARS-CoV-2 test was 100.0% (13/13) in the preliminary and final result reporting phases in sensitivity cohort 1, where the nasopharyngeal swab specimens were suspended directly into the mariPOC sample buffer or first into saline (Table 1). Detailed description of the positive samples are shown in supplemental material (Online Resource). Prevalence of SARS-CoV-2 in the first sample cohort was 6%, which is well in alignment with the prevalence during the study time in the geographical area (5%).

**Table 1.** Sensitivity of the mariPOC SARS-CoV-2 test when compared with the qRT-PCR methods.

| Sample cohort | mariPOC result phase | No. of specimens |  | Sensitivity (%) (95% CI) |
| --- | --- | --- | --- | --- |
|  |  | TP | FN |  |
| 1 | Final | 13 | 0 | 100.0 (75.3– 100.0) |
|  | Preliminary | 13 | 0 | 100.0 (75.3– 100.0) |
| 2 | Final | 38 | 7 | 84.4 (70.5–93.5) |
|  | Preliminary | 33 | 12 | 73.3 (58.1–85.4) |

TP = true positive, FN = false negative, CI = confidence interval (exact Clopper-Pearson method).

The sensitivity of the test in sensitivity cohort 2 was 73.3% (33/45) and 84.4% (38/45) in the preliminary and final result reporting phases, respectively, when the nasopharyngeal swabs were initially suspended in undefined transport mediums and further diluted with the mariPOC sample buffer (Table 1). Based on 95% confidence interval, both cohorts had similar statistical reliability (Table 1). Overall, 38 out of 45 samples were positive with the mariPOC in sensitivity cohort 2 (Fig. 2). The test showed 100% (31/31) positivity rate compared to qRT-PCR for Ct values ≤28 (Table 2). Above the Ct value 28, the positivity rate of the mariPOC declined as typical for an antigen test, reaching 91.9% (34/37) with Ct values ≤30.

**Table 2.**
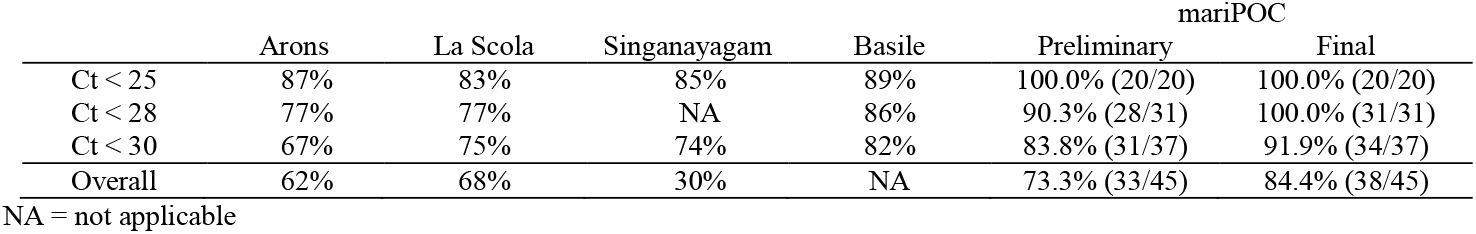
**Comparison of** cumulative positivity rates of viral culture (four studies) and mariPOC (sensitivity sample cohort 2) to qRT-PCR below different Ct categories. (10, 12, 14, 47)

**Fig. 2.**
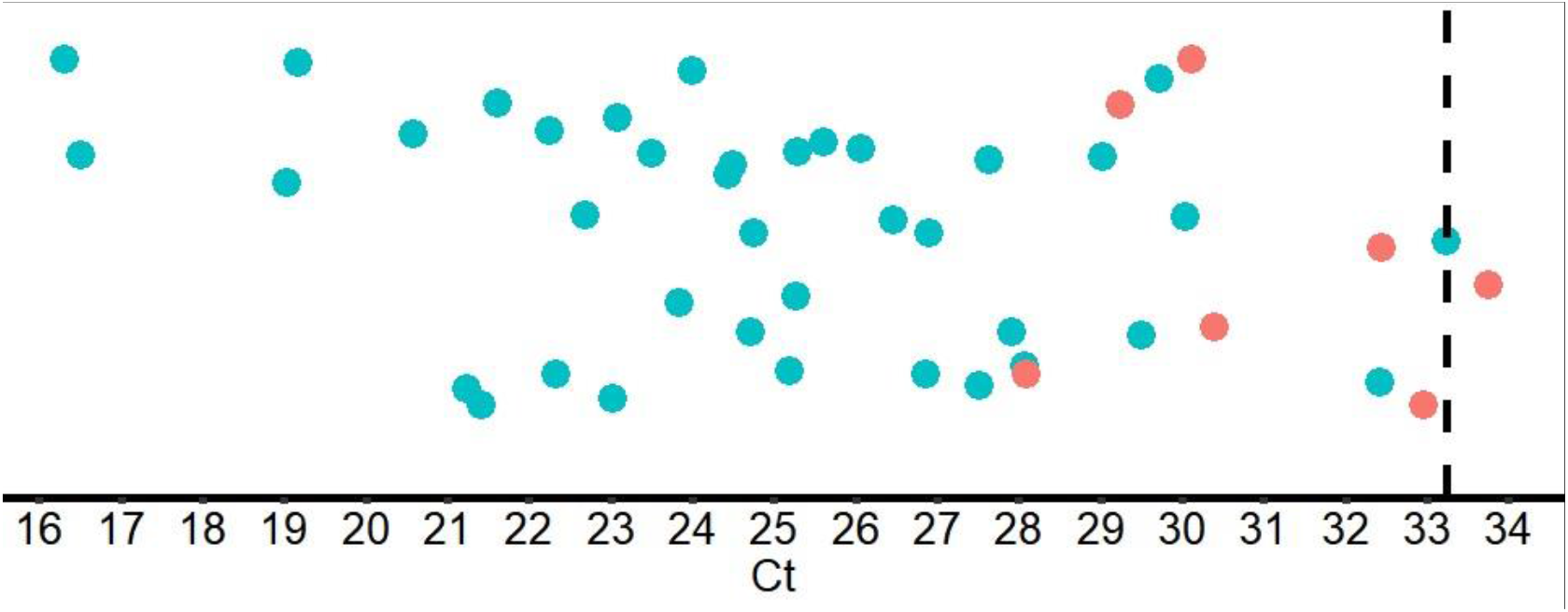
Ct values of qRT-PCR for mariPOC test positive (green dots) and negative (red dots) samples in the validation sample cohort 2. Dashed line is at Ct 33.24, which was the lowest detected Ct.

## Discussion

When setting up a diagnostic process or choosing a diagnostic method, one should carefully consider, to start with, whether the disease, clinical condition and use case, require high sensitivity for ruling out or high specificity for ruling in. There is a need for both in fighting the COVID-19 pandemic. In general, analytically highly sensitive testing, such as PCR testing, is good at ruling out a disease (e.g. keeping a ward clean) while highly specific testing, such as antigen testing, is good at ruling in a disease (e.g. acute infection diagnostics and assessing infectiousness). Because of rapidity and lesser logistic challenges compared to central lab testing, antigen testing is particularly good in surveillance, field-testing, screening of masses, cohorting of inpatients, acute disease diagnostics, and in assessing the infectiousness of individuals (13, 38). Especially when disease prevalence is low, clinical specificity of the screening and diagnostic testing should be emphasized to keep unnecessary quarantines and economic damages at minimum while still allowing sufficient enough infection control (29).

According to scientific data, to effectively prevent spread of the disease, pandemic control should prioritize accessibility, frequency of testing, and rapid sample-to-answer time over test sensitivity (37, 39, 40). Viral load and probability to infect others is highest just prior to onset of symptoms and during the symptomatic phase (41).

We describe here analytical and clinical validation of mariPOC SARS-CoV-2 test sensitivity and specificity. Determination of LoD was performed with gamma-irradiation inactivated viral culture supernatant and showed that only less than three infectious units per test was needed for positive test result. LoD was also determined as qRT-PCR Ct units using UV-inactivated virus (Online Resource). The obtained Ct LoD was 33, which approaches the theoretical analytical sensitivity of a typical PCR method with 5 µl cDNA volume and applying 95% confidence interval. The maximum Ct values detected in clinical samples (Fig. 2 and Table 2.) were similar to the determined Ct LoD.

Based on high identity (89.1%) between SARS-CoV-1 (Uniprot entry, P59595) and SARS-CoV-2 (UniProt entry, P0DTC9) nucleocapsid protein sequences, and obtaining positive result for purified SARS-CoV-1 nucleocapsid protein in cross-reactivity testing (Table S1 in the Online Resource), it is highly likely that the mariPOC test detects also the SARS-CoV-1 virus itself.

Cross-reactions were not observed. A minor limitation of the study is that cross-reactivity for MERS coronavirus and coronavirus HKU1 were assessed using purified protein and sequence analysis (Online Resource) and not with clinical samples or cultured virus.

Our sensitivity validation cohort 1 showed 100% sensitivity. While the highest qRT-PCR Ct obtained by the primary reference test in this cohort was 30.2, the cohort consisted of unselected and consecutive samples collected from patients with clear symptoms. This might explain why there were no samples with higher Ct values. The sensitivity cohort 2 showed 100% positivity rate for the mariPOC below qRT-PCR Ct 28 (Table 2 and Fig. 2). This result was excellent taking into account that the samples were unfavorable for the mariPOC platform with separation-free fluorescent measurement. Colorful transport media are not recommended for mariPOC testing since they elevate fluorescent signal levels (42) and unnecessarily dilute the samples, which reduces sensitivity. The pooled sensitivity of sample cohorts 1 and 2 at qRT-PCR Ct ≤ 30 was 94%, suggesting even higher sensitivity for mariPOC compared to what has been reported in the literature for SARS-CoV-2 viral culture against qRT-PCR, as summarized in Table 2. In addition, our results are in line with at least two other N-protein detecting tests that were evaluated against RT-PCR and culture (43, 44). Several studies have shown that infectivity of SARS-CoV-2 declines rapidly in samples showing qRT-PCR Ct above 25, and viable virus is rarely isolated after 8 days from onset of the symptoms. The detection of sole viral RNA, especially at low levels without the detectable level of viral N-protein or culture positivity, is a questionable marker of acute infection and infectiousness (10-14, 16, 45-47).

The results suggest that the clinical sensitivity of the mariPOC test (84.4% in unfavorable sample matrix to 100.0% when used according to manufacturer recommendations) is similar or even better than that of at least some rapid RT-PCR tests (93.4%), (37) when symptomatic pati ents suspected with acute COVID-19 infection are tested within the first five days of symptoms and prevalence among tested samples is reasonable (6% in sensitivity study 1). Recommended sample in the mariPOC test is native nasopharyngeal swab specimen suspended into 1.3 mL of the RTI sample buffer. Other specimen types may yield in lower apparent sensitivity. In the sensitivity cohort 1, suspending part of the swabs first into saline prior to the addition of mariPOC RTI sample buffer followed by a further dilution into mariPOC RTI sample buffer by a factor of two for the testing, diluted the specimens 4 to 20 times (2 to 4.3 PCR Ct units) more than the recommended sample pretreatment. Additional dilution lowers the sensitivity compared to the recommended protocol, and could have led to an underestimation of the test sensitivity. Strengths of the sensitivity validation included that the specimens were collected in the early phase of the COVID-19 pandemic in Finland that minimized the detection of RNA persistence with the RT-PCR among the cohort population.

Limitations of the validation study were that the patient characteristics and the number of symptomatic days before sampling were not available for the study. Freezing and thawing of the positive samples prior to mariPOC testing and additional dilutions to the recommended protocol were also limitations. However, if any, these could have had a negative effect on the mariPOC test sensitivity and, hence, the study at least did not overestimate the sensitivity of the mariPOC SARS-CoV-2 test.

## Conclusions

The mariPOC SARS-CoV-2 test is an automated, highly specific and clinically accurate test with rapid sample-to-answer time for individuals with clinical suspicion and in acute phase of an infection. The closed tube test system and the design of operational steps minimize specimen handling and possible exposure of user to infectious material. The multianalyte syndromic tests help to differentiate between SARS-CoV-2 and other viruses, such as influenza. The single analyte test provides high capacity of 300 samples a day at the point-of-sampling. Objective result read-out and LIS connectability minimize manual work and human errors. Our study together with other scientific data suggests that the mariPOC can detect majority of the cases already in 20 minutes with sensitivity similar to a rapid PCR while maximum sensitivity is achieved in 55 minutes. The positivity rate of mariPOC compared to qRT-PCR Ct values in clinical samples is very high up to Ct 28-30, and samples at least up to Ct 33.24 (Fig. 2) or 35.78 (Online Resourcetable 2) can be detected depending on the PCR method and gene target. The detection of conserved epitope in the N-protein of SARS coronaviruses with the mariPOC likely provides accurate information about infectiousness similarly to other antigen tests and viral culture and suggests ability to detect also emerging virus variants. Further studies using viral culture as comparative method and follow-up of infectiousness of patients using antigen detection are needed in order to optimize viral respiratory tract infection management.

## Supporting information

Online Resource Supplemental material

## Data Availability

Anonymised line data is available upon request.

## Supplemental material

Supplemental material is available as Online Resource. Online Resource1, PDF file, 0.4 MB.

## Acknowledgements

We thank Vita laboratories Ltd and Seoul Clinical Laboratory for performing RT-PCR analyses and Titta Lehtonen performing the mariPOC analyses in SataDiag.

## Compliance with ethical standards

## Author contributions

JMK and PA: Major contributions in the development of the mariPOC test, in scientific design and execution of the studies, result analysis, scientific analysis, and writing of the manuscript.

KH: Major contributions in providing specimens for clinical validation, in scientific analysis and revising the manuscript.

AH: Major contributions in providing specimens for clinical validation, in scientific analysis and revising the manuscript.

NI and CS-K: Major contributions in scientific analysis, and revision of the manuscript.

KS: Major contributions in providing specimens for clinical validation, in scientific analysis and revising the manuscript.

JOK: Major contributions in the development of the mariPOC test and in scientific design, execution, and analysis of the results, and revision of the manuscript.

