## Supplementary material for "Clinical Validation of Automated and Rapid mariPOC SARS-CoV-2 Antigen Test": Online Resource Supplemental material

#### ANALYTICAL SPECIFICITY

Cross-reactions of the mariPOC® SARS-CoV-2 test were studied against different respiratory tract infection causing pathogens and normal flora microbes. Viral and bacterial samples were favored instead of recombinant antigen if they were reasonably available.

Viral concentrations were verified to be high, based on quantitative results obtained using the corresponding analyte specific mariPOC® tests (Europe, IVD CE) at ArcDia's Research and Development laboratory. The mariPOC® tests are qualitative for the end user, but the technology on which the mariPOC® is based on, ArcDia™ two-photon excitation assay technique, is quantitative in nature. For seasonal coronaviruses 229E and NL63, high viral load in culture supernatant was verified with non-commercial mariPOC® research reagents detecting these viruses.

Cross-reactions of the mariPOC® tests with the coronavirus types SARS-CoV-1, SARS-CoV-2 and MERS-CoV were studied by analyzing 1000 ng/mL in reaction concentrations of purified nucleocapsid proteins. Concentration of $4x10^7$ bct/ml (OD600 = 0.04) in reaction, assumed to be high positive, was used for bacterial cross-reactivity testing. Bacterial concentration determination was based on assumption that OD600 = 1.0 corresponds to $10^9$ bct/ml.

According to the results, (Table S1) mariPOC® SARS-CoV-2 test did not cross-react with the tested microbes. Recognition of SARS-CoV-1 was expected based on the high similarity in N-protein sequence between the two SARS coronaviruses. Cross-reactivity testing for MERS coronavirus (UniProt entry, K9N4V7) and coronavirus HKU1 (UniProt entry, Q5MQC6) was possible only using purified protein. It is highly unlikely that the mariPOC® test would cross-react with either of these viruses as they share only 44.2% (UniProt entry, K9N4V7) and 28.9% identity (UniProt entry, Q5MQC6), respectively, in their nucleocapsid protein peptide sequence with the SARS-CoV-2.

**Supplementary table 1** Cross-reaction study information and results.

| Analyzed sample | Source | Test concentration | Test result (+ or -) |
|---|---|---|---|
| Human coronavirus OC43 (purified virus preparation) | ATCC | 5x dilution from viral culture supernatant | - |
| Human coronavirus strain 229E (purified virus preparation) | ATCC | 5x dilution from viral culture supernatant | - |
| Human coronavirus strain NL63 (purified virus preparation) | ATCC | 5x dilution from viral culture supernatant | - |
| SARS coronavirus 1 (purified nucleoprotein) | ArcDia International Ltd, Finland | 1 µg/mL | + |

| Analyzed sample | Source | Test concentration | Test result (+ or -) |
|---|---|---|---|
| MERS coronavirus (purified nucleoprotein) | ArcDia International Ltd, Finland | 1 µg/mL | - |
| Human coronavirus strain HKU1 (recombinant nucleoprotein) | ArcDia International Ltd, Finland | 10 µg/mL | - |
| Influenza A virus (purified virus preparation): H3N2: A/Panama/2007/99 | Hytest Ltd, Turku, Finland | 5 µg/mL | - |
| Influenza A virus (purified virus preparation): H3N2: A/Texas/50/12 H3N2: A/Victoria/361/11 | Research Institute of Influenza, St Petersburg, Russia | 10 µg/mL | - |
| Influenza A virus (purified virus preparation): H1N1: A/New Caledonia/20/99 | Biomarket Ltd, Turku Finland | 5 µg/mL | - |
| Influenza A virus (so-called swine flu, supernatant of virus culture): H1N1v: A/FIN/554/09 | Finnish institute for health and welfare | 40x dilution from viral culture supernatant | - |
| Influenza B virus (purified virus preparation): Phuket/3073/2013 | Research Institute of Influenza, St Petersburg, Russia | 100x dilution from viral culture supernatant | - |
| Respiratory syncytial virus (purified virus preparation): Type A / Long | AbD Serotec Inc, now Bio-Rad Laboratories, Inc | 7.4x dilution from viral culture supernatant | - |
| Respiratory syncytial virus (purified virus preparation): Type B / clinical strain (Labquality round 1, 2017) | Labquality Ltd, Helsinki, Finland | 2x dilution from viral culture supernatant | - |
| Human metapneumovirus (supernatant of virus culture) | Department of Virology, University of Turku, Finland | 10x dilution | - |
| Human bocavirus (clinical specimen) | Turku University Hospital, Finland | Nasopharyngeal swab suspended in 1.3 mL of sample buffer. High positive in specific antigen test | - |
| Human bocavirus (recombinant VP2 antigen) | Vilnius University, Institute of Biotechnology, Lithuania | 5 µg/mL | - |
| Parainfluenza 1 virus (purified virus preparation): Sendai | Hytest Ltd, Turku, Finland | 50 µg/mL | - |
| Parainfluenza 2 virus (purified virus preparation) | Virology department, University of Turku, Finland | 25x dilution from viral culture supernatant | - |
| Parainfluenza 3 virus (purified virus preparation): Washington/1957 C243 | AbD Serotec Inc, now Bio-Rad Laboratories, Inc | 20 µg/mL | - |
| Adenovirus strain 6 (purified virus preparation) | Hytest Ltd, Turku, Finland | 4 µg/mL | - |
| *Streptococcus pneumoniae* (bacterial culture suspension inactivated by heating) ATCC 49619 | Finnish institute for health and welfare | $4 \times 10^7$ bacteria/mL | - |
| Group A streptococci / *S. pyogenes*: ATCC 19615 (bacterial culture suspension inactivated by heating) | Finnish institute for health and welfare | $4 \times 10^7$ bacteria/mL | - |
| *Staph. aureus* (bacterial culture suspension inactivated by heating): ATCC 29213 | Finnish institute for health and welfare | $4 \times 10^7$ bacteria/mL | - |
| *Staph. epidermidis* (isolate, bacterial culture | Finnish institute for health | $4 \times 10^7$ bacteria/mL | - |

| Analyzed sample | Source | Test concentration | Test result (+ or -) |
|---|---|---|---|
| suspension inactivated by heating) | and welfare | | |
| *S. anginosus species* (isolates)*:*<br>*S. anginosus* (209), *S. constellatus* (5690) and *S. intermedius* (1343) (bacterial culture suspension inactivated by heating). | Finnish institute for health and welfare | 4 x $10^7$ bacteria/mL | - |
| *H. influenzae* (bacterial culture suspension inactivated by heating): ATCC 33391 | Finnish institute for health and welfare | 4 x $10^7$ bacteria/mL | - |
| *H. parainfluenzae* (bacterial culture suspension inactivated by heating): ATCC 33392 | Finnish institute for health and welfare | 4 x $10^7$ bacteria/mL | - |

#### CLINICAL SENSITIVITY, SAMPLE COHORT 1

Out of 211 specimens primary comparator SARS-CoV-2 RT-PCRs (Allplex™ and Amplidiag®) reported 13 positive samples (Table S2). One sample (ID 106) was not originally tested with the comparator RT-PCR for an unknown reason, but this sample was positive in the confirmatory PCR. Hence, altogether 14 PCR positive samples were reported. Out of 14 PCR positive samples, mariPOC® SARS-CoV-2 test gave a positive result for 13 samples. Comparator RT-PCR, thus, reported one additional positive samples (number 197) compared to mariPOC®. However, confirmatory RT-PCR from swab samples taken for mariPOC® testing was negative in both cases for all gene targets. Sample ID 197 was considered as false positive in comparator RT-PCR test as it was negative with the confirmatory PCR from the same saline sample used for mariPOC® testing. The conclusion was supported by the fact that the original comparator RT-PCR showed Ct values (on average 33.4) higher than the theoretical 95% CI sensitivity limit of qRT-PCR.

**Supplementary table 2.** mariPOC® and RT-PCR SARS-CoV-2 line data results for samples giving positive result in one or more tests.

| Sample number | Sample type | Result | | | |
|---|---|---|---|---|---|
| | | mariPOC® | | Comparator RT-PCR (Gene, Ct value) | Confirmatory RT-PCR* (Gene, Ct value) |
| | | Preliminary | Final | | |
| 5 | Dry swab | + | + | + (NA) | NA |
| 7 | Dry swab | + | + | + (NA) | NA |
| 13 | Dry swab | + | + | + (NA) | - All genes neg |
| 21 | Dry swab | + | + | + (NA) | NA |
| 47 | Dry swab | + | + | + (NA) | NA |
| 62 | Dry swab | + | + | + (NA) | NA |
| 95 | Swab in saline | + | + | + (NA) | NA |
| 106 | Swab in saline | + | + | NA# | + (E, 30.2; RdRP, 35.2; N 35.78) |
| 151 | Dry swab | + | + | + (NA) | NA |
| 197 | Swab in saline | - | - | + (N, 35.6; ORF1ab, 31.16) | - All genes neg |
| 198 | Swab in saline | + | + | + (N, 21.89; ORF1ab, 20.66) | NA |
| 202 | Swab in saline | + | + | + (N, 23.58; ORF1ab, 21.8) | NA |
| 203 | Swab in saline | + | + | + (N, 28.82; ORF1ab, 26.5) | NA |
| 206 | Swab in saline | + | + | + (N, 24.41; ORF1ab, 24.15) | NA |

NA, Not available/applicable.

* Three samples were analyzed with confirmatory RT-PCR

### Sample was not originally tested with comparator RT-PCR for an unknown reason. For the sensitivity comparison of mariPOC®, the primary comparator RT-PCR was assumed to have been given a positive result. It was considered statistically unlikely that the comparator PCR would have given a false negative result for exactly the specimen for, which it was not done for an unknown reason related to routine testing process.

**UV-INACTIVATED VIRUS**

Preparation of the UV-inactivated SARS-CoV-2 culture supernatant ($2 \times 10^6$ PFU/mL, Ct 17, University of Helsinki, Finland) is described in the referenced study. The LoD for this preparation was determined similarly as for the gamma-inactivated virus.

Rusanen J, Kareinen L, Szirovicza L, Uğurlu H, Levanov L, Jääskeläinen A, Ahava M, Kurkela S, Saksela K, Hedman K, Vapalahti O, Hepojoki J. 2020. A generic, scalable, and rapid TR-FRET –based assay for SARS-CoV-2 antigen detection. medRxiv https://doi.org/10.1101/2020.12.07.20245167.
